# Representation of evidence-based clinical practice guideline recommendations on FHIR

**DOI:** 10.1101/2022.05.16.22275120

**Authors:** Gregor Lichtner, Carlo Jurth, Brian S Alper, Claudia Spies, Martin Boeker, Joerg J Meerpohl, Falk von Dincklage

## Abstract

**Background:** Various formalisms have been developed to represent clinical practice guideline recommendations in a computer-interpretable way. However, none of the existing formalisms leverage the structured and computable information that emerge from the evidence-based guideline development process. Thus, we here propose a FHIR-based guideline representation format that is structurally aligned to the knowledge artifacts emerging during the process of evidence-based guideline development.

**Methods:** We identified the information required to represent evidence-based clinical practice guideline recommendations and reviewed the knowledge artifacts emerging during the evidence-based guideline development process. Then we conducted a consensus-based design process with domain experts to develop an information model for guideline recommendation representation that is structurally aligned to the evidence-based guideline recommendation development process and a corresponding representation based on evidence-based medicine (EBM)-on-FHIR resources.

**Results:** The information model of clinical practice guideline recommendations and its EBMonFHIR-based representation contain the clinical contents of individual guideline recommendations, a set of metadata for the recommendations, the ratings for the recommendations (e.g., strength of recommendation, certainty of overall evidence), the ratings of certainty of evidence for individual outcomes (e.g., risk of bias) and links to the underlying evidence (systematic reviews based on primary studies). We created profiles and an implementation guide for all FHIR resources required to represent a complete clinical practice guideline and used the profiles to implement an exemplary clinical guideline recommendation.

**Conclusions:** Our EBMonFHIR-based representation of clinical practice guideline recommendations allows to directly link the evidence assessment process through systematic reviews and evidence grading, and the underlying evidence from primary studies to the resulting guideline recommendations. This not only allows to evaluate the evidence on which recommendations are based on transparently and critically, but also allows for a more direct and in future automatable way to generate computer-interpretable guideline recommendations based on computable evidence.

## Introduction

A plethora of approaches have been developed to specify computer-interpretable representations of clinical practice guideline recommendations, such as Asbru, EON, GLIF3, SAGE or GUIDE [1–7]. Despite their many differences, all these representations have in common that they are designed based on the concept that computer-interpretable guideline recommendations are derived by translating unstructured recommendations into the formalism of the respective representation. However, with the emergence of evidence-based medicine over the last decades [8], guideline recommendations are developed in a structured process in which knowledge artifacts, such as the effect size estimates from primary studies or the grading of available evidence, are derived at each step. Leveraging these structured knowledge artifacts for the computer-interpretable representation of clinical guideline recommendations might require a reconsideration of the current computer-interpretable guideline recommendation formalisms.

The systematic development process of evidence-based guideline recommendations for a specific clinical question is based on a systematic review of studies providing data that address the clinical question, potentially followed by aggregations of the study data in meta-analyses and finally balancing and grading all available information in structured evidence-to-decision frameworks, resulting in trustworthy recommendations [9–12]. With digitalization spreading, more and more structured data becomes available from this process. Considering that this structured data is the basis to formulate unstructured guideline recommendations, translating these unstructured guideline recommendations back into structured recommendation representations induces unnecessary workload. As an alternative approach, we propose a format to represent computer-interpretable guideline recommendations in a structure that directly builds on the knowledge artifacts emerging during the guideline recommendation development process.

To represent such knowledge artifacts that emerge during guideline recommendation development, the HL7 Clinical Decision Support (CDS) Work Group-sponsored Evidence-based Medicine (EBM) on Fast Healthcare Interoperable Resources (FHIR) project sub-work group (EBMonFHIR) created a collection of FHIR resources [13]. These FHIR resources provide a standardized way of describing data formats and elements that are related to both the evidence generation and evidence assessment parts of evidence-based guideline development, such as the effect size for certain outcomes of a specific intervention on a specific population, the grading of the certainty of evidence, and metadata such as the group of authors or publication status and version of knowledge artifacts.

Based on these resources, we developed an interoperable, computer-interpretable representation of clinical practice guideline recommendations that combines the structured data emerging during the development of evidence-based guideline recommendations, from the evidence generation in primary studies and systematic reviews and the structured evidence assessment in evidence-to-decision frameworks to the resulting guideline recommendations.

## Methods

### Requirements Engineering

To establish which information about the contents and the metadata of clinical practice guideline recommendations need to be captured in the FHIR-based representation, we performed an iterative consensus-based requirements engineering process. In this process, we included five clinical stakeholders (health care professionals) and five guideline developers from German university hospitals, medical societies and Cochrane Germany to identify both the required information for practical use of clinical guideline recommendations and the metadata that is required to assess e.g. the credibility and strength of recommendations, as well as the information required to connect individual recommendations to their underlying evidence from systematic reviews of primary studies. Participants of the requirements engineering process were recruited from the members of the COVID-19 evidence ecosystem (CEOsys) project of the German COVID-19 Research Network of University Medicine (“Netzwerk Universitätsmedizin”) [14].

### Information Modelling

After establishing the list of required information, we developed an information model of the collected items by identification of the item types, item cardinalities, relationships between items and grouping of the items. Input from clinical stakeholders and guideline developers helped defining the entity relationships for the items. The Cochrane PICO Ontology guided discussions for medical relationships among the recommendation contents [15]. The candidate models were reviewed with clinical stakeholders and medical information scientists for semantic and syntactic appropriateness and completeness.

### FHIR Mapping

Following the definition of the information model, we performed an iterative consensus-based process to map the model’s items and relationships to their correspondences in FHIR. We included FHIR developers and EBMonFHIR maintainers as well as the clinical stakeholders and guideline developers from the requirements engineering process for necessary feedback and validation of the mapping.

In close collaboration with the EBMonFHIR maintainers, we derived a mapping of the information model to existing EBMonFHIR resources. During this process, suboptimal applicability of the resources to our use case were resolved by introducing differential changes to the EBMonFHIR resources in the FHIR specification. We therefore developed our FHIR profiles and implementation guide based on the latest available FHIR development build, which was at the time of writing the daily continuous integration build of FHIR R5 as of April 1, 2022.

### Development

The specification of the appropriate use of FHIR resources for representation of clinical guideline recommendations was performed by means of creating FHIR resource profiles and an implementation guide. Development was performed using software versioning and continuous integration and continuous development (CI/CD) workflows on GitHub [16].

To maximize ease of implementation and adherence to the FHIR standard, the development of FHIR profiles was carried out under the condition that no extensions should be used in the profiles unless necessary.

Profiling was performed using the FHIR ShortHand (FSH) language (version 1.2.0) and the SUSHI software (version 2.3.0) for translation to FHIR structure definitions in JSON format. For each profile, we required that at least one instance (i.e., example) was defined that instantiated that profile. Automated syntax and code checking were performed using the HL7 FHIR validator as implemented in the FSH validator python package (version 0.2.2; [17]). This package was also used to test and validate each defined instance against the profile it instantiates in the CI/CD workflow.

The implementation guide was created using the HL7 FHIR IG Publisher tool version 1.1.115-SNAPSHOT (modified) and the FHIR core artifacts version 5.6.41-SNAPSHOT (modified). Both software tools were modified to implement the continuous integration FHIR build as of April 1, 2022. Modified sources and compiled executables are provided in the respective repositories of our GitHub organization [18].

A custom template was used for creation of the implementation guide’s html pages. The creation and deployment of the implementation guide to GitHub pages [19] is automatically triggered by commits to the main branch of the project’s GitHub repository in order to always keep the source profiles and the implementation guide in synchronization.

### Evaluation

To evaluate syntactic and semantic completeness and appropriateness of the EBMonFHIR-based guideline recommendation representation, we implemented a recent COVID-19 guideline recommendation for the treatment of hospitalized patients. The implementation was reviewed with clinical stakeholders and medical information scientists to confirm the appropriateness and completeness of the implemented information.

## Results

### Information model

We identified the required and optional information that need to be captured in a representation of clinical guideline recommendations and categorized these information items into groups on two dimensions (Figure 1). The first dimension describes the content type categories of the information items including (i) items describing the medical relationships, (ii) the metadata items such as study types or authors, and (iii) items describing the justification for the recommendation, including statistics of primary studies and systematic reviews as well as the gradings of individual outcomes and of the final guideline recommendation. The second dimension describes the stages of the guideline recommendation development process from (i) the primary evidence generation in clinical studies, (ii) the systematic review and evidence assessment of individual outcomes, and (iii) the derivation of the recommendation including evidence grading across all outcomes. In the following sections, we provide details on each of the content type categories.

**Figure 1.**
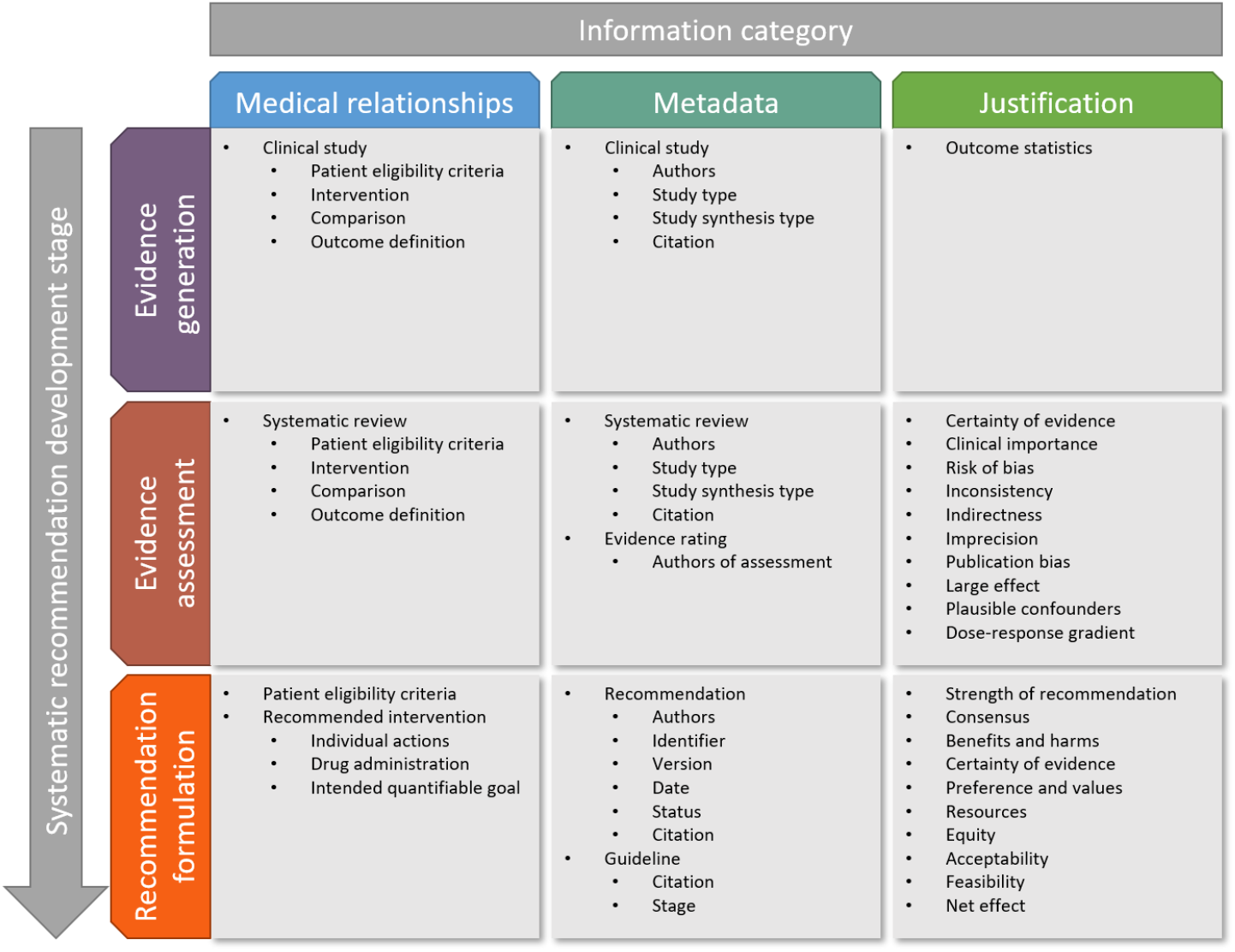
High level information model of the guideline recommendation representation.

### Medical relationships

The medical relationships that a clinical question is made of can be decomposed using the “PICO” framework into the four components of Population, Intervention, Comparison and Outcome. These components distinctly define to which patients a clinical question applies (population), which intervention or treatment is considered (intervention), which control treatment is used as a comparison for the investigated intervention (comparison) and which outcome is considered (outcome). Using the PICO framework to decompose the clinical questions of primary clinical studies and meta-analyses is standard methodology in evidence-based medicine [20]. To leverage the PICO framework to describe the medical relationships of guideline recommendations, only the two components P and I are required. These allow to define distinctly in which patients (the “population”) which treatment or action (the “intervention”) is recommended. As guideline recommendations are commonly defined as absolute recommendations and not in relation to a specific comparison, the comparison component of PICO is not required to describe the medical relationship of a guideline recommendation. Likewise, as guideline recommendations summarize across all outcomes, the individual outcomes are not a required component of the recommendation itself. Instead, a guideline recommendation could include an “outcome net effect” (see “justification” below), but this does not require describing a specific outcome on the guideline recommendation level.

### Metadata

The metadata category considers descriptive metadata such as the authors of all parts of the guideline development process including those of the clinical studies, the systematic review, the evidence assessment, and of the grading of the final recommendation. The metadata may also include versioning information of the recommendation (e.g., version number, date, publication status), which are particularly important in the context of living guidelines that are regularly updated. Additionally, citations of individual studies, published systematic reviews, recommendations and guidelines are part of the metadata.

### Justification

The justification category considers all information items on the basis of which the recommendation is ultimately given. For individual outcomes, these include the effect size (i.e., outcome statistics) of the intervention as determined from clinical studies or the systematic review and furthermore the quality of evidence rating according to GRADE [12]. For the final recommendation as a whole, the justification includes ratings according to the GRADE Evidence-to-Decision (EtD) [9,10] as well as the outcome net effect of the recommended intervention (weighting of benefits and harms) [21].

### FHIR Mapping

#### Overview

The structure of our EBMonFHIR-based guideline representation combines multiple interlinked FHIR resources for the representation of the different parts and aspects of a clinical practice guideline and the individual recommendations (Table 1). The definition of the recommended intervention serves as the root of the structure and references the group of patients (“population”) for which the intervention is recommended, as well as the recommendation ratings according to the GRADE EtD framework (Figure 2). We created profiles for each resource to consistently constrain the representation of the clinical guideline recommendations. The following sections describe each of the profiles in more detail.

**Table 1.** Information items of evidence-based clinical practice guidelines and their mapping to FHIR. Shown are the items and the FHIR resource name and profile name, which are used to represent the respective item.

| Category | Item | FHIR Resource(s) | Profile Name(s) | Required |
| --- | --- | --- | --- | --- |
| Guideline | Collection of recommendations | Composition | Guideline |  |
| Recommendation | Population | EvidenceVariable | RecommendationEligibilityCriteria | ✓ |
|  | Intervention | PlanDefinition,<br>ActivityDefinition | RecommendationPlan,<br>RecommendationAction | ✓ |
|  | Recommendation Citation | Citation | RecommendationCitation |  |
|  | Guideline Citation | Citation | GuidelineCitation |  |
| Recommendation Justification | Strength of Recommendation | ArtifactAssessment | RecommendationJustification |  |
|  | Consensus |  |  |  |
|  | Benefits and Harms |  |  |  |
|  | Certainty of Evidence |  |  |  |
|  | Preference and values |  |  |  |
|  | Resources |  |  |  |
|  | Equity |  |  |  |
|  | Acceptability |  |  |  |
|  | Feasibility |  |  |  |
| Outcome | Outcome Definition | EvidenceVariable | OutcomeDefinition |  |
|  | Certainty of Evidence | ArtifactAssessment | CertaintyOfEvidenceRating |  |
|  | Clinical Importance |  |  |  |
|  | Risk of Bias |  |  |  |
|  | Inconsistency |  |  |  |
|  | Indirectness |  |  |  |
|  | Imprecision |  |  |  |
|  | Publication Bias |  |  |  |
|  | Large Effect |  |  |  |
|  | Plausible Confounders |  |  |  |
|  | Dose-Response Gradient |  |  |  |
| Net Effect | Net Effect | Evidence | NetEffectEstimate |  |
| Studies (primary & systematic reviews) | Intended Study Population | EvidenceVariable | StudyEligibilityCriteria |  |
|  | Intervention/Comparison | EvidenceVariable | InterventionDefinition |  |
|  | Study Citation | Citation | StudyCitation |  |
| Primary Studies | Observed Study Population | Group | StudyCohort |  |
|  | Outcome Statistics (relative effect / mean difference) | Evidence | StudyOutcomeEvidence |  |
| Systematic reviews | Observed Populations from Primary Studies | Group | EvidenceSynthesisCohorts |  |
|  | Aggregated Outcome Statistics | Evidence | OutcomeEvidenceSynthesis |  |
|  | Observed Outcomes from Primary Studies | EvidenceVariable | EvidenceDataSet |  |

**Figure 2.**
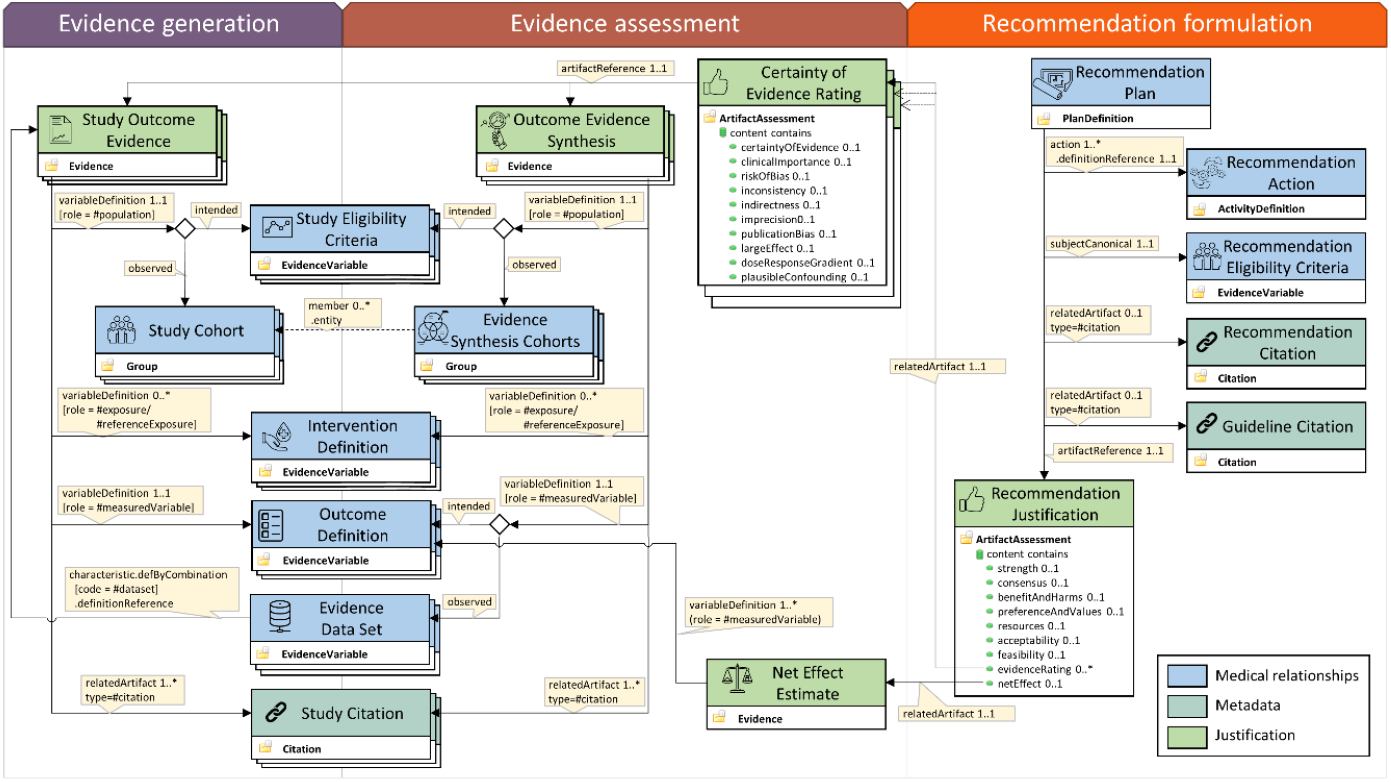
Overview of the resources and profiles used for representing clinical practice guideline recommendations on EBMonFHIR. Note that depending on the type of study (primary study vs systematic review), the PICO elements (eligibility criteria, intervention definition, outcome definition) for the evidence may be part of evidence generation or evidence assessment, respectively.

#### Intervention

The recommended intervention is represented in the *RecommendationPlan* profile of the *PlanDefinition* resource. This resource allows to specify detailed clinical workflows, consisting of individual actions that are to be taken (or should not be taken) during specified circumstances. These individual actions are defined using the *RecommendationAction* profile of the *ActivityDefinition* resource and are referenced by the *RecommendationPlan* profile.

The *RecommendationPlan* profile serves as the root of an individual guideline recommendation representation, referencing – directly or indirectly – all further defining parts of an individual guideline recommendation. Most importantly, the profile directly references the definition of the population, for which the specified intervention is recommended, via the *subjectReference* field. The *RecommendationPlan* profile, together with the *RecommendationAction* profile and the *RecommendationEligibilityCriteria* profiles form the required core of a computer-interpretable guideline recommendation representation that allows for automated integration with clinical data to provide clinical decision support.

Next to the core components, the *RecommendationPlan* profile references via the *relatedArtifact* field the *RecommendationJustification* profile of the *ArtifactAssessment* resource to represent the justification of the recommendation in accordance with the GRADE EtD framework. Additionally, the *RecommendationPlan* profile references the *RecommendationCitation* and *GuidelineCitation* profiles of the *Citation* resource to provide references to the published recommendation and/or guideline that contains the recommendation, respectively.

The *InterventionDefinition* profile of *EvidenceVariable* is used to represent the general, non-executable definition of an intervention in clinical studies or systematic reviews. In contrast to *RecommendationPlan* and *RecommendationAction*, the intervention represented by the *InterventionDefinition* profile does not usually describe the intervention in sufficient detail to provide automated clinical decision support.

#### Population

The patients to which the recommended intervention defined in the *RecommendationPlan* profile is applicable is specified via the *RecommendationEligibilityCriteria* profile of the *EvidenceVariable* resource. This profile leverages the newly introduced option of the *EvidenceVariable* resource to specify complex characteristics for a population definition, such as temporal dependencies between individual characteristics. Thereby, the *EvidenceVariable* resource allows to specify much more complex population characteristics than the *Group* resource, and such complex definitions are required to represent population definitions from actual guideline recommendations as well as from study eligibility criteria.

#### Recommendation Justification

To represent the rating of the recommendation regarding its strength, the achieved consensus among the guideline developers, the certainty of evidence and additional ratings according to the GRADE EtD framework [10], the *RecommendationJustification* profile of the *ArtifactAssessment* resource is used. The *ArtifactAssessment* resource is newly introduced in FHIR Release 5 and is used to represent one or more comments, ratings, or classifications about another resource. Here, that other resource is the *RecommendationPlan* profile that describes the guideline recommendation. The profile includes the individual ratings as justification for the recommendation in the content field and uses different value sets and code systems for the individual components (Table 2).

**Table 2.** Ratings in the RecommendationJustification profile. Shown are the ratings that are represented in the RecommendationJustification profile, together with the value set binding on content.classifier, the code system used in the respective value set, the organization that defined the code system and the codes included in the respective value set.

| Item | ValueSet | CodeSystem | Defined by | Codes |
| --- | --- | --- | --- | --- |
| **Strength of Recommendation** | RecommendationStrength | RecommendationStrength | CEOsyst | strong-for \| strong-against \| weak-for \| weak-against |
| **Consensus** | RatingConsensus | StrengthOfRecommendationRating | FHIR | strong \| weak |
| **Benefits and Harms** | RatingCertaintyOfEvidence | EvidenceCertaintyRating | FHIR | high \| moderate \| low \| very-low |
| **Certainty of Evidence** | RatingBenefitAndHarms | EvidenceToDecisionCertaintyRating | CEOsyst | small-net-benefit \| substantial-net-benefit \| important-harms |
| **Preference and values** | RatingPreferenceAndValues | EvidenceToDecisionCertaintyRating | CEOsyst | factor-not-considered \| substantial-variability \| no-substantial-variability \| few-want-intervention |
| **Resources** | RatingResources | EvidenceToDecisionCertaintyRating | CEOsyst | factor-not-considered \| important-issues-or-not-investigated \| no-important-issues \| important-negative-issues |
| **Equity** | RatingEquity | EvidenceToDecisionCertaintyRating | CEOsyst | factor-not-considered \| important-issues-or-not-investigated \| no-important-issues \| intervention-increases-inequity |
| **Acceptability** | RatingAcceptability | EvidenceToDecisionCertaintyRating | CEOsyst | factor-not-considered \| important-issues-or-not-investigated \| no-important-issues \| intervention-poorly-accepted |
| **Feasibility** | RatingFeasibility | EvidenceToDecisionCertaintyRating | CEOsyst | factor-not-considered \| important-issues-or-not-investigated \| no-important-issues \| intervention-difficult-to-implement |

Additionally, *RecommendationJustification* references the ratings of individual evidence (i.e., primary studies or meta-analyses) in accordance with GRADE via the *CertaintyOfEvidenceRating* profile of *ArtifactAssessment*. The *RecommendationJustification* profile also references the *NetEffectEstimate* profile of the Evidence resource to describe the combined net effect (net harm / net benefit) of the recommendation given the considered outcomes [21].

A single instance of the *RecommendationJustification* profile is used for a particular recommendation.

#### Certainty of Evidence Rating

The ratings of the individual outcomes that are relevant to a specific recommendation are represented using the *CertaintyOfEvidenceRating* profile of the *ArtifactAssessment* resource (Table 3). The assessed artifact for each *CertaintyOfEvidenceRating* instance is an instance of the *StudyOutcomeEvidence* or *OutcomeEvidenceSynthesis* profile of the *Evidence* resource, which represents the statistics related to that specific outcome and references the systematically collated body of evidence from which the effect estimates are derived. For each individual outcome, a different instance of the *CertaintyOfEvidenceRating* profile is used.

**Table 3.** Ratings in the CertaintyOfEvidenceRating profile. Shown are the ratings that are represented in the CertaintyOfEvidenceRating profile, together with the value set binding on content.classifier, the code system used in the respective value set, the organization that defined the code system and the codes included in the respective value set.

| Item | ValueSet | CodeSystem | Defined by | Codes |
| --- | --- | --- | --- | --- |
| **Certainty of Evidence** | RatingCertaintyOfEvidence | EvidenceCertaintyRating | FHIR | high \| moderate \| low \| very-low |
| **Clinical Importance** | RatingClinicalImportance | ClinicalImportance | CEOsyst | 1 \| 2 \| 3 \| 4 \| 5 \| 6 \| 7 \| 8 \| 9 |
| **Risk of Bias** | RatingConcernDegree | EvidenceCertaintyRating | FHIR | no-concern \| serious-concern \| very-serious-concern \| extremely-serious-concern |
| **Inconsistency** | RatingConcernDegree | EvidenceCertaintyRating | FHIR | no-concern \| serious-concern \| very-serious-concern \| extremely-serious-concern |
| **Indirectness** | RatingConcernDegree | EvidenceCertaintyRating | FHIR | no-concern \| serious-concern \| very-serious-concern \| extremely-serious-concern |
| **Imprecision** | RatingConcernDegree | EvidenceCertaintyRating | FHIR | no-concern \| serious-concern \| very-serious-concern \| extremely-serious-concern |
| **Publication Bias** | RatingConcernDegree | EvidenceCertaintyRating | FHIR | no-concern \| serious-concern \| very-serious-concern \| extremely-serious-concern |
| **Large Effect** | RatingUpratingTwoLevels | EvidenceCertaintyRating | FHIR | no-change \| upcode1 \| upcode2 |
| **Plausible Confounders** | RatingUpratingOneLevel | EvidenceCertaintyRating | FHIR | no-change \| upcode1 |
| **Dose-Response Gradient** | RatingUpratingOneLevel | EvidenceCertaintyRating | FHIR | no-change \| upcode1 |

#### Outcome

The definition of an individual outcome (e.g., 30-day all-cause mortality), possibly including parameters such as timing and method of outcome measurement, is represented using the *OutcomeDefinition* profile of *EvidenceVariable*.

The evidence for individual outcomes, i.e., summary statistics from systematic reviews of clinical studies or from primary studies, are represented by an instance of the *StudyOutcomeEvidence* profile (for primary studies) or *OutcomeEvidenceSynthesis* (for systematic reviews) of the *Evidence* resource. These instances hold the statistics associated with the particular outcome (e.g., the relative risk for intervention vs. comparison and the associated 95% confidence interval). For that purpose, *StudyOutcomeEvidence* and *OutcomeEvidenceSynthesis* may reference to the definition of an intervention and a comparison via the *InterventionDefinition* profile of *EvidenceVariable*.

As the study population may be different from the recommendation population, the *StudyOutcomeEvidence* and *OutcomeEvidenceSynthesis* profiles references the *StudyEligibilityCriteria* profile of *EvidenceVariable* for the *intended* definition of the population in the studies from which the evidence is derived. For the actually *observed* study population, the *StudyCohort* profile of *Group* is used for primary studies to describe the number of participants, and the *EvidenceSynthesisCohorts* profile of *Group* is used for systematic reviews, describing the total number studies and referencing the individual *StudyCohort* instances from the primary studies.

As the evidence on which a systematic review is based on is a dataset of results from primary studies, the *OutcomeEvidenceSynthesis* references the *EvidenceDataSet* profile of *EvidenceVariable*, which combines the *StudyOutcomeEvidence* instances from the primary studies into the dataset of the systematic review.

Additionally, *StudyOutcomeEvidence* and *OutcomeEvidenceSynthesis* may reference *StudyCitation* instances for the published systematic review and/or primary studies.

#### Net Effect

The net effect is the overall expected effect of following a recommendation, calculated as an importance-weighted average of the individual outcome effects. It thus quantifies the balance between desirable and undesirable effects of an intervention [21]. When the net effect should be specified, it can be represented using the *NetEffectEstimate* profile of the *Evidence* resource. It references the individual outcomes that are included in the calculation of the net effect and their relative importance weightings as model characteristics.

#### Guidelines

To represent the collection of multiple recommendations in a guideline, the *Guideline* profile of the *Composition* resource is used. The *Composition* resource sections are references to the individual *RecommendationPlan* instances that define the individual recommendations. Additionally, the guideline publication may be referenced via the *GuidelineCitation* profile of the *Citation* resource.

The whole collection of all resources defined here may be bundled using the *GuidelineBundle* profile of the *Bundle* resource.

### Terminologies

To identify the different contents of clinical practice guideline recommendations according to the PICO framework, we used concepts from the Cochrane PICO ontology [15] as well as SNOMED CT concepts if available. To identify the rating types and values of the evidence-to-decision framework, we used code systems defined by HL7 FHIR and developed new code systems if required (Table 2, Table 3). To represent the clinical variables, the vocabularies intended to be used are the *Systematized Nomenclature of Medicine - Clinical Terms* (SNOMED CT) for conditions, substances, and general concepts [22], *Logical Observation Identifiers Names and Codes* (LOINC) for laboratory observation [23], *International Statistical Classification of Diseases and Related Health Problems, 10*^*th*^ *revision* (ICD-10) for diagnoses [24], *Anatomical Therapeutic Chemical Classification System* (ATC) for drugs [25] and *Unified Code for Units of Measure* (UCUM) for measurement units [26]. Other vocabularies may be used when suitable.

### Evaluation

To validate our proposed guideline recommendation structure, we implemented a recent recommendation from a guideline for the treatment of COVID-19 intensive care patients [27–29]. As this recommendation is part of a living guideline, it is ideally suited to validate the agility of our proposed guideline recommendation representation as new evidence is added or certainty ratings for specific outcomes or the actual recommendations changed. We selected a recommendation that included both a treatment that should be performed on a specific population of patients and one that should not be performed on another population of patients. Specifically, the guideline includes a recommendation for treating ventilated COVID-19 patients with the systemic corticosteroid Dexamethasone, applied for 10 days orally or intravenously, and a recommendation against treating non-ventilated patients with any Dexamethasone. The recommendation is based on a systematic review [30], which is part of the evidence generation process. The different outcomes, most importantly all-cause mortality, was evaluated as part of the evidence assessment process and the recommendations were formulated based on the evaluated evidence (Figure 3). We created instances for all appropriate profiles and included them as examples in the implementation guide [19] and on the FEvIR platform [31].

**Figure 3.**
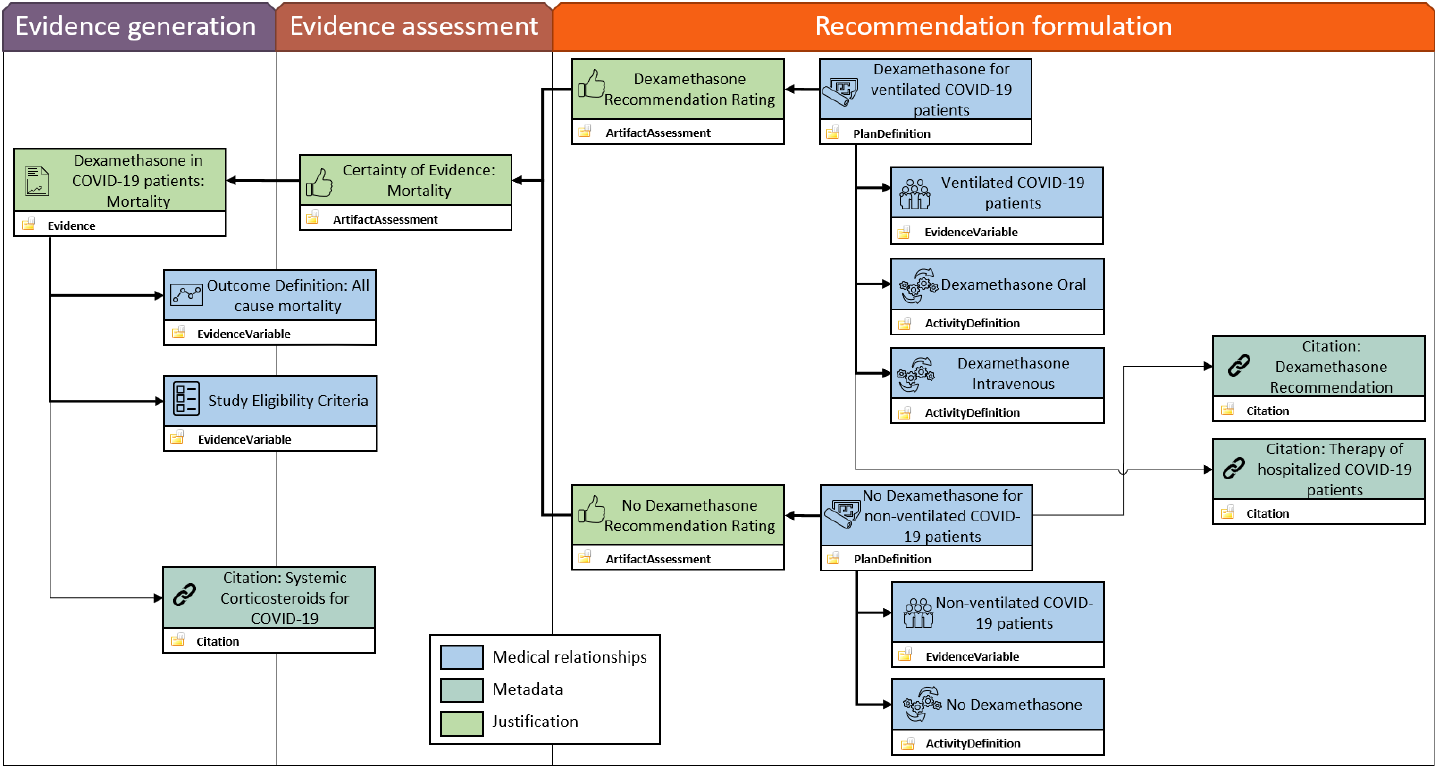
Overview of the guideline recommendation implementation from a treatment guideline for hospitalized COVID-19 patients.

## Discussion

We here present an EBMonFHIR-based representation for computer-interpretable guideline recommendations that leverages the structured data emerging during the development of evidence-based guidelines, ranging from primary evidence-generating studies over systematic reviews and the evidence assessment using evidence-to-decision frameworks to the formulated recommendation. We have developed the representation based on an iterative, consensus-based requirements engineering process and mapping of the thereby derived information model items to EBMonFHIR resources. We developed profiles for all used FHIR resources and created an implementation guide for the guideline representation format. To evaluate the format, we implemented a recent guideline recommendation in our representation format.

The here presented computer-interpretable representation of the whole evidence-based guideline development process offers a set of advantages over just representing the final guideline recommendations:

First, representing the evidence from primary studies and systematic reviews (i.e., effect size statistics) and evidence-to-decision process artifacts in a computer-interpretable way allows them to be used for semi-automated guideline recommendation formulation. Indeed, knowledge artifacts from evidence generation and evidence assessment (i.e., ratings of existing evidence) in our representation format may be published in repositories, e.g. on collaborative guideline development platforms such as MAGICapp [32], and be reused for different guideline recommendation development processes.

Second, the representation of the evidence generation and evidence assessment artifacts may be particularly valuable during the lifecycle of living guidelines that are regularly updated as new evidence is published or existing evidence is re-evaluated in light of new findings.

Third, integrating a complete guideline recommendation representation into clinical decision support or recommendation monitoring systems allows to close the loop from evidence to recommendation back to evidence: Currently, the information flow is unidirectional from evidence generation via evidence assessment to the formulated recommendation. However, when these recommendations are implemented in a hospital setting by means of automated integration with electronic health records (EHR), the treatment of patients according or not according to the recommendation in connection with appropriate outcome data generates real-world evidence for (or against) the intervention. This evidence, readily representable in the here proposed guideline recommendation representation format, can be evaluated and used in an update process of the recommendation.

Fourth, representation of the full process of evidence-based recommendation development allows the target audience of the guideline recommendations a direct and transparent assessment of the evidence generation and evidence assessment process that underlies the recommendations. It has to be noted that the structured evidence generation and evidence assessment artifacts are generated in any case within the context of a structured guideline recommendation development process; representing them in a computer-interpretable way therefore comes at no or only little further cost during guideline recommendation development.

Even though some of the existing guideline recommendation formalisms that only represent the final guideline recommendations would allow to represent all of the information, the effort to newly encode it from scratch appears as a relevant barrier. In contrast, the focus of our formalism is to enable using data generated during the guideline recommendation development process, thus reducing the required effort to include the additional data. However, as each representation formalism has its own advantages and specialized scope of functions, our proposed representation might complement existing formalisms instead of substituting them. In that way, mapping the treatment recommendation part of our representation format to other formalisms would allow to close the gap between these formalisms and the knowledge artifacts emerging during evidence-based guideline recommendation development.

Apart from the previously described differences in content, in contrast to most previous guideline recommendation formalisms, we have based our representation on FHIR, which currently might be considered the most important standard for defining interoperable medical data exchange, with growing support from EHR software vendors. There has been considerable work done on representing the contents of clinical guideline recommendations in FHIR, namely the CPG-on-FHIR project [33]. This project, like the EBMonFHIR project sponsored by the HL7 Clinical Decision Support Work Group, has developed a comprehensive implementation guide for the representation of guideline recommendation contents. However, in contrast to our project, the CPG-on-FHIR implementation guide does not include profiles for linking the full evidence generation and evidence assessment process to the final guideline recommendations – although for example the strength of recommendation and quality of evidence are supported via FHIR extensions. Additionally, the CPG-on-FHIR project uses the *Group* resource for the definition of the population part of guideline recommendations, whereas we are using the newly introduced characteristics backbone element of the *EvidenceVariable* resource, which allows for much more complex definitions of patient group characteristics than does the *Group* resource. To allow more complex definitions, the use of clinical quality language (CQL) was introduced into the *Group* resource recently; additionally, it would be possible to outsource the population characteristics definition into newly introduced concepts from a code system. However, both solutions outsource the definition into another language or domain, potentially reducing interoperability and limiting leveraging the query capabilities that are offered by pure FHIR-based solutions. However, the CPG-on-FHIR project and our guideline recommendation representation format are not mutually exclusive and combining efforts in the future may increase the utility of both approaches.

In the current work, we have not restricted the use of concepts in the definition of the population or intervention to particular code systems such as SNOMED CT or LOINC. However, a major aim of our representation, as of any computer-interpretable guideline recommendation formalism, is the automated applicability in computerized clinical decision support systems. In such a setting, the concepts used for the definition of the population and intervention must be mapped to the concepts used in individual EHR systems in clinics. To reduce the need for additional mapping of concept and variables, a possible approach is to provide the clinical data from the EHR system in the OMOP common data model (CDM), as this uses a standardized vocabulary for clinical concepts, and further constraining the profiles for the intervention (*RecommendationPlan, RecommendationAction*) and population (*RecommendationEligibilityCriteria*) to only allow concepts from the OMOP standardized vocabulary. While this shifts the need of mapping individual EHR systems to concepts to the mapping to the OMOP CDM, the provision of clinical data in OMOP CDM allows for a large range of clinical and research use case in the individual clinics, far beyond the integration of clinical guideline recommendations with the clinical data. However, constraining FHIR resources to the use of OMOP standardized vocabulary concepts is currently hampered by the unavailability of that vocabulary as a FHIR *CodeSystem* resource and additional work is required to better link the two domains of FHIR and OMOP regarding OMOP’s standard vocabulary [34].

Our proposed FHIR-based representation of guideline recommendations has three main limitations: First, it is based on EBMonFHIR resources, which are mostly at a low maturity level and subject to frequent, even breaking, changes. The implementation guide therefore needs to be constantly kept in synchronization with current developments of EBMonFHIR resources. However, this is ensured by our active collaboration and participation in the development of the EBMonFHIR resources.

Second, there is currently no execution engine available for our representation format that would allow to automatically integrate the recommended interventions with clinical data to provide clinical decision support. However, a prototype implementation for such an execution engine to be used with clinical data in the OMOP common data model (CDM) format is currently being developed. Additionally, translators may be implemented that translate the FHIR-based representation to other guideline recommendation formalisms that already have an execution engine implemented (e.g. the GLIF3 execution engine or the SAGE execution engine for EON [35,36]).

Third, not all information for guideline recommendation execution can be expressed in current FHIR resources: The dependence and relationship between recommendations from the same or different guidelines cannot be modeled in the *PlanDefinition* resource without the introduction of extensions.

We expect that in the maturation process of resources from the Clinical Decision Support group, which maintains the *PlanDefinition* resource, and EBMonFHIR, additional concepts and relationships of declarative process modelling will be introduced to completely represent relationships between recommendations and guidelines.

The proposed EBMonFHIR-based guideline recommendation representation is currently being implemented in several German university hospitals, where it is used as a computer-interpretable guideline recommendation representation to be automatically integrated with standardized clinical patient data to provide information about individual patient guideline recommendation applicability and adherence. In the context of this project, and as the EBMonFHIR resources evolve, our guideline recommendation representation will be continuously updated and improved where necessary.

## Conclusion

To leverage the structured, computable knowledge artifacts that emerge during evidence-based guideline recommendation development, we have developed a FHIR-based guideline recommendation representation that is aligned with these knowledge artifacts. Thereby, our EBMonFHIR-based representation of clinical practice guideline recommendations allows to directly link the systematic evidence assessment and the underlying evidence from systematic reviews and primary studies to the resulting guideline recommendations. This not only allows for a transparent and critical evaluation of the evidence on which recommendations are based, but also provides a more straightforward and, in the future, automatable way to generate computer-interpretable guideline recommendations from the available evidence.

## Data Availability

All data produced are available online at https://github.com/ceosys/cpg-on-ebm-on-fhir

## Code availability

All code used for the development of the profiles and the implementation guide is available at https://github.com/CEOsys/cpg-on-ebm-on-fhir. The implementation guide is hosted on https://ceosys.github.io/cpg-on-ebm-on-fhir/. All code systems, value sets and examples are additionally hosted on the FEvIR platform [31].

## Supplementary Figures

**Supplementary Figure 1.**
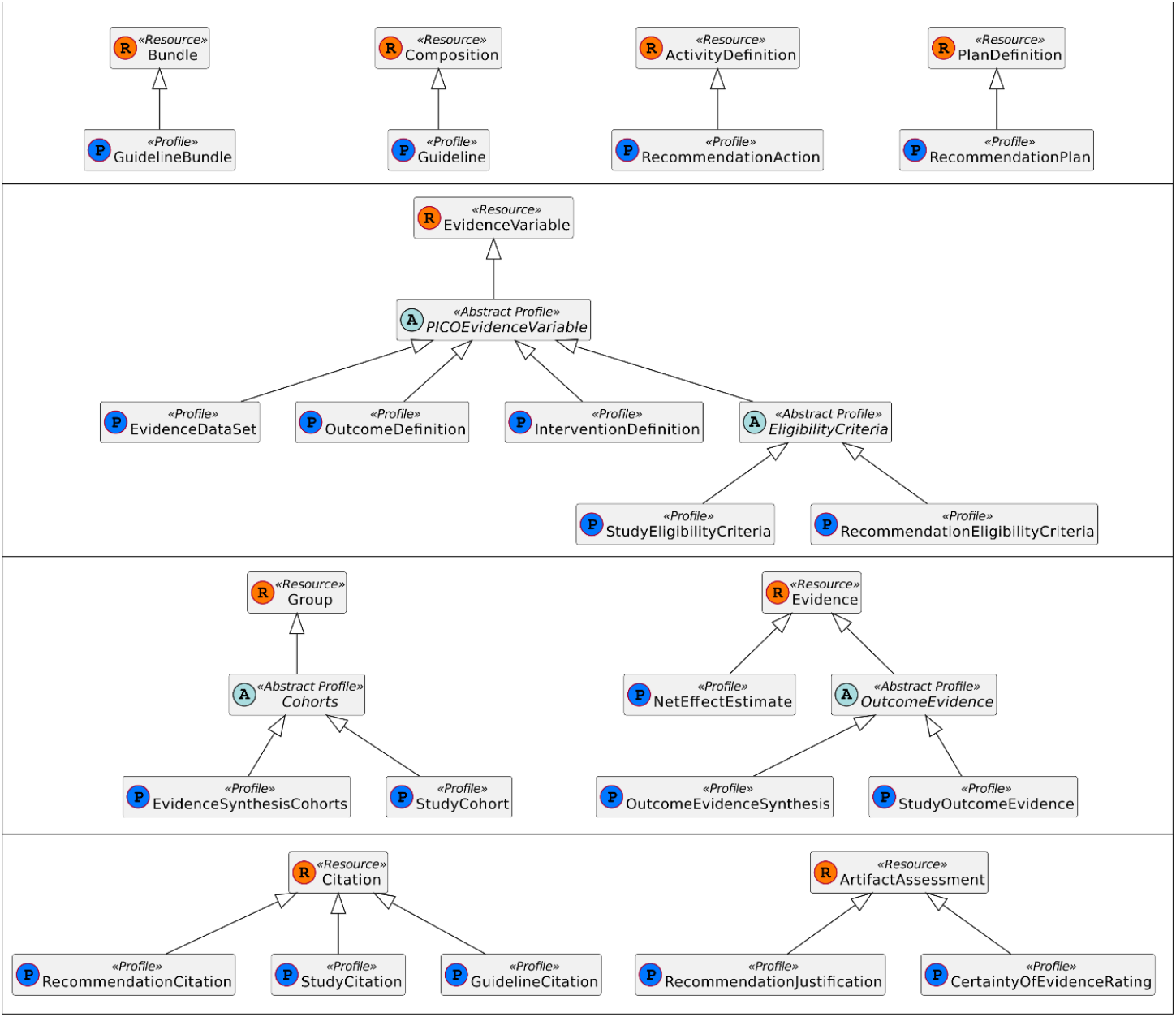
Class Diagram of CPG-on-EBMonFHIR profiles.

## References

[1] A. Seyfang, S. Miksch, M. Marcos, Combining diagnosis and treatment using asbru, Int. J. Med. Inf. 68 (2002) 49–57. https://doi.org/10.1016/S1386-5056(02)00064-3.

[2] S.W. Tu, M.A. Musen, Modeling Data and Knowledge in the EON Guideline Architecture, MEDINFO 2001. (2001) 280–284. https://doi.org/10.3233/978-1-60750-928-8-280.

[3] A.A. Boxwala, M. Peleg, S. Tu, O. Ogunyemi, Q.T. Zeng, D. Wang, V.L. Patel, R.A. Greenes, E.H. Shortliffe, GLIF3: a representation format for sharable computer-interpretable clinical practice guidelines, J. Biomed. Inform. 37 (2004) 147–161. https://doi.org/10.1016/j.jbi.2004.04.002.

[4] S.W. Tu, J.R. Campbell, J. Glasgow, M.A. Nyman, R. McClure, J. McClay, C. Parker, K.M. Hrabak, D. Berg, T. Weida, J.G. Mansfield, M.A. Musen, R.M. Abarbanel, The SAGE Guideline Model: Achievements and Overview, J. Am. Med. Inform. Assoc. JAMIA. 14 (2007) 589–598. https://doi.org/10.1197/jamia.M2399.

[5] P. Ciccarese, E. Caffi, S. Quaglini, M. Stefanelli, Architectures and tools for innovative Health Information Systems: The Guide Project, Int. J. Med. Inf. 74 (2005) 553–562. https://doi.org/10.1016/j.ijmedinf.2005.02.001.

[6] M. Peleg, Computer-interpretable clinical guidelines: A methodological review, J. Biomed. Inform. 46 (2013) 744–763. https://doi.org/10.1016/j.jbi.2013.06.009.

[7] D. Riaño, M. Peleg, A. ten Teije, Ten years of knowledge representation for health care (2009– 2018): Topics, trends, and challenges, Artif. Intell. Med. 100 (2019) 101713. https://doi.org/10.1016/j.artmed.2019.101713.

[8] B. Djulbegovic, G.H. Guyatt, Progress in evidence-based medicine: a quarter century on, The Lancet. 390 (2017) 415–423. https://doi.org/10.1016/S0140-6736(16)31592-6.

[9] P. Alonso-Coello, H.J. Schünemann, J. Moberg, R. Brignardello-Petersen, E.A. Akl, M. Davoli, S. Treweek, R.A. Mustafa, G. Rada, S. Rosenbaum, A. Morelli, G.H. Guyatt, A.D. Oxman, the G.W. Group, GRADE Evidence to Decision (EtD) frameworks: a systematic and transparent approach to making well informed healthcare choices. 1: Introduction, BMJ. 353 (2016) i2016. https://doi.org/10.1136/bmj.i2016.

[10] P. Alonso-Coello, A.D. Oxman, J. Moberg, R. Brignardello-Petersen, E.A. Akl, M. Davoli, S. Treweek, R.A. Mustafa, P.O. Vandvik, J. Meerpohl, G.H. Guyatt, H.J. Schünemann, the G.W. Group, GRADE Evidence to Decision (EtD) frameworks: a systematic and transparent approach to making well informed healthcare choices. 2: Clinical practice guidelines, BMJ. 353 (2016) i2089. https://doi.org/10.1136/bmj.i2089.

[11] R. Graham, M. Mancher, D. Miller Wolman, S. Greenfield, E. Steinberg, eds., Clinical Practice Guidelines We Can Trust, National Academies Press (US), Washington (DC), 2011. https://doi.org/10.17226/13058.

[12] G. Guyatt, A.D. Oxman, E.A. Akl, R. Kunz, G. Vist, J. Brozek, S. Norris, Y. Falck-Ytter, P. Glasziou, H. deBeer, R. Jaeschke, D. Rind, J. Meerpohl, P. Dahm, H.J. Schünemann, GRADE guidelines: 1. Introduction—GRADE evidence profiles and summary of findings tables, J. Clin. Epidemiol. 64 (2011) 383–394. https://doi.org/10.1016/j.jclinepi.2010.04.026.

[13] B. Alper, M. Mayer, K. Shahin, J. Richardson, L. Schilling, M. Tristan, N. Salas, Achieving evidence interoperability in the computer age: setting evidence on FHIR, BMJ Evid.-Based Med. 24 (2019) A15–A15. https://doi.org/10.1136/bmjebm-2019-EBMLive.28.

[14] What is CEOsys?, CEOsys. (n.d.). https://covid-evidenz.de/what-is-ceosys/ (accessed April 8, 2022).

[15] Cochrane PICO Ontology, (n.d.). https://data.cochrane.org/ontologies/pico/index-en.html (accessed March 15, 2022).

[16] Clinical Practice Guidelines on EBMonFHIR, (2022). https://github.com/CEOsys/cpg-on-ebm-on-fhir (accessed March 15, 2022).

[17] G. Lichtner, FHIR Shorthand Validator, (2021). https://github.com/glichtner/fsh-validator (accessed March 15, 2022).

[18] CEOsys, GitHub. (n.d.). https://github.com/CEOsys (accessed March 15, 2022).

[19] Clinical Practice Guidelines (CPG) on EBMonFHIR, (n.d.). https://ceosys.github.io/cpg-on-ebm-on-fhir/ (accessed March 15, 2022).

[20] C. Schardt, M.B. Adams, T. Owens, S. Keitz, P. Fontelo, Utilization of the PICO framework to improve searching PubMed for clinical questions, BMC Med. Inform. Decis. Mak. 7 (2007) 16. https://doi.org/10.1186/1472-6947-7-16.

[21] B.S. Alper, P. Oettgen, I. Kunnamo, A. Iorio, M.T. Ansari, M.H. Murad, J.J. Meerpohl, A. Qaseem, M. Hultcrantz, H.J. Schünemann, G. Guyatt, Defining certainty of net benefit: a GRADE concept paper, BMJ Open. 9 (2019) e027445. https://doi.org/10.1136/bmjopen-2018-027445.

[22] Snomed CT, SNOMED. (n.d.). https://www.snomed.org/ (accessed March 16, 2022).

[23] C.J. McDonald, S.M. Huff, J.G. Suico, G. Hill, D. Leavelle, R. Aller, A. Forrey, K. Mercer, G. DeMoor, J. Hook, W. Williams, J. Case, P. Maloney, for the Laboratory LOINC Developers, LOINC, a Universal Standard for Identifying Laboratory Observations: A 5-Year Update, Clin. Chem. 49 (2003) 624–633. https://doi.org/10.1373/49.4.624.

[24] International Classification of Diseases (ICD), (n.d.). https://www.who.int/standards/classifications/classification-of-diseases (accessed March 22, 2022).

[25] Anatomical Therapeutic Chemical (ATC) Classification, (n.d.). https://www.who.int/tools/atc-ddd-toolkit/atc-classification (accessed March 22, 2022).

[26] The Unified Code for Units of Measure, (n.d.). https://ucum.org/trac (accessed March 16, 2022).

[27] S. Kluge, U. Janssens, T. Welte, S. Weber-Carstens, G. Schälte, Christoph D. Spinner, Jakob J. Malin, Petra Gastmeier, Florian Langer, Martin Wepler, Michael Westhoff, Michael Pfeifer, Klaus F. Rabe, Florian Hoffmann Bernd W. Böttiger, Julia Weinmann-Menke, Alexander Kersten, Peter Berlit, Marcin Krawczyk, Wiebke Nehls, Reiner Haase, Monika Nothacker, Gernot Marx, Christian Karagiannidis, S3-Leitlinie Empfehlungen zur stationären Therapie von Patienten mit COVID-19, AWMF (Arbeitsgemeinschaft der Wissenschaftlichen Medizinischen Fachgesellschaften e.V.), 2021. https://www.awmf.org/leitlinien/detail/ll/113-001LG.html.

[28] S. Kluge, U. Janssens, C.D. Spinner, M. Pfeifer, G. Marx, C. Karagiannidis, Guideline group, Clinical Practice Guideline: Recommendations on Inpatient Treatment of Patients with COVID-19, Dtsch. Arzteblatt Int. 118 (2021) arztebl.m2021.0110. https://doi.org/10.3238/arztebl.m2021.0110.

[29] J.J. Malin, C.D. Spinner, U. Janssens, T. Welte, S. Weber-Carstens, G. Schälte, P. Gastmeier, F. Langer, M. Wepler, M. Westhoff, M. Pfeifer, K.F. Rabe, F. Hoffmann, B.W. Böttiger, J. Weinmann-Menke, A. Kersten, P. Berlit, M. Krawczyk, W. Nehls, F. Fichtner, S. Laudi, M. Stegemann, N. Skoetz, M. Nothacker, G. Marx, C. Karagiannidis, S. Kluge, Key summary of German national treatment guidance for hospitalized COVID-19 patients : Key pharmacologic recommendations from a national German living guideline using an Evidence to Decision Framework (last updated 17.05.2021), Infection. (2021). https://doi.org/10.1007/s15010-021-01645-2.

[30] C. Wagner, M. Griesel, A. Mikolajewska, A. Mueller, M. Nothacker, K. Kley, M.-I. Metzendorf, A.-L. Fischer, M. Kopp, M. Stegemann, N. Skoetz, F. Fichtner, Systemic corticosteroids for the treatment of COVID-19, Cochrane Database Syst. Rev. 8 (2021) CD014963. https://doi.org/10.1002/14651858.CD014963.

[31] FEvIR Project - CPG-on-EBMonFHIR, FEvIR Platf. (n.d.). https://fevir.net/resources/Project/46952 (accessed May 13, 2022).

[32] MAGICapp, (n.d.). https://app.magicapp.org/ (accessed March 21, 2022).

[33] FHIR Clinical Guidelines (v1.0.0) (STU 1), (n.d.). http://hl7.org/fhir/uv/cpg/index.html (accessed March 15, 2022).

[34] M. Lawley, J. Steel, J. Grimes, FHIR Terminology Services for OMOP – opportunities report, (n.d.) 21.

[35] D. Wang, M. Peleg, S.W. Tu, A.A. Boxwala, O. Ogunyemi, Q. Zeng, R.A. Greenes, V.L. Patel, E.H. Shortliffe, Design and implementation of the GLIF3 guideline execution engine, J. Biomed. Inform. 37 (2004) 305–318. https://doi.org/10.1016/j.jbi.2004.06.002.

[36] D. Isern, A. Moreno, Computer-based execution of clinical guidelines: A review, Int. J. Med. Inf. 77 (2008) 787–808. https://doi.org/10.1016/j.ijmedinf.2008.05.010.

